# What it takes to implement AI in Africa: health-system lessons from developing an ML-enabled maternal risk stratification algorithm in Tanzania

**DOI:** 10.64898/2026.07.26.26358974

**Authors:** Augustino Hellar, Isaac Lyatuu, Alen Kinyina, Yusuph Kulindwa, Edwin Ernest, Hamid Mandali, Cyprian Mtani, Husna Athumani, Frank Phiri, Presley Massawe, Omari Sukari, Phineas Sospeter, Ntuli Kapologwe

## Abstract

**Background:** Machine learning (ML) has growing potential to support early identification of high-risk pregnancies in resource-constrained settings. However, most studies focus on model development and predictive performance, with less attention to the health-system processes required to generate ML-ready data and translate risk information into clinical action. The Mlinde Mama Project in Tanzania combined Group Antenatal Care (G-ANC), digital maternal health systems, and development of an ML-enabled risk stratification model for hypertensive disorders of pregnancy (HDP). This study examined the health-system conditions shaping the pathway from routine care to actionable ML-enabled risk information.

**Methods:** We conducted a retrospective mixed-methods implementation analysis of this project that was implemented between 2022 and 2024 in Geita, Tanzania. The analysis triangulated endline evaluation findings, quantitative exit interviews with pregnant women, focus group discussions with women and healthcare workers, key informant interviews, project implementation records, routine data from Tanzania’s Unified Community System (UCS), and documented ML development experience. Relevant quotations from the final evaluation report were systematically screened, selected, and coded. An abductive analysis combined inductively derived themes with the Non-adoption, Abandonment, Scale-up, Spread and Sustainability (NASSS) framework and a study-specific ML implementation pathway.

**Results:** Seven themes emerged across three phases of the implementation pathway. ML readiness began before the algorithm, with reliable clinical measurement and documentation; patient participation and task-sharing redistributed data generation. The paper-to-digital transition shaped which information became ML-ready data. The potential value of ML depended on workflow redesign rather than prediction alone. Technology created an efficiency paradox in which task-sharing reduced workload while staffing shortages and confirmatory work generated new burdens. Advanced analytics depended on basic infrastructure, while sustainability relied on teamwork, trust, user demand, and local ownership.

**Conclusions:** Our findings suggest that successful ML implementation in maternal health begins with health-system readiness before the algorithm itself. A critical paper-to-digital transition stage was a major determinant of data quality and ML readiness. ML-enabled maternal risk stratification should therefore be approached as a sociotechnical intervention spanning measurement, digitization, clinical confirmation, and follow-up. This implementation readiness is critical for the successful development of an ML-enabled risk stratification system. Crucially, our findings resonate with the key domains of the NASSS framework, underscoring how addressing multidimensional complexities, from technological design to organizational readiness, is vital for successful adoption, scale-up and long-term sustainability.

**Author Summary:** Machine learning (ML) has shown considerable potential for improving maternal health by identifying women at increased risk of complications such as hypertensive disorders of pregnancy. However, most studies focus on how accurately ML models predict risk, with less attention paid to the health-system conditions required for these technologies to function in routine care, particularly in low-resource settings.

We examined implementation lessons from the Mlinde Mama Project in Tanzania, which combined Group Antenatal Care, digital health tools and ML-enabled maternal risk stratification across six public health facilities. We triangulated qualitative and quantitative evaluation findings, project implementation records and routine digital health data to understand how clinical information moves from routine care to digital systems and, ultimately, to clinical action.

We found that ML readiness begins long before an algorithm is deployed. Reliable clinical measurements, successful conversion of paper records into digital data, workflow design, workforce capacity, infrastructure and trust all influenced whether risk information could become useful in practice. Our study highlights a critical stage in the context of low-resource settings, a “paper-to-digital transition” stage, where information may be delayed, duplicated or lost before reaching an ML system.

These findings suggest that ML should be implemented as a sociotechnical intervention rather than as a standalone technology.

## 1. Introduction

Hypertensive disorders of pregnancy (HDP) remain a major cause of maternal morbidity and mortality globally and disproportionately affect women in low- and middle-income countries, accounting for roughly 14% of maternal deaths worldwide[1]. In Tanzania, HDP contributes substantially to direct obstetric mortality; at Muhimbili National Hospital, severe pre-eclampsia and eclampsia accounted for 19.9% of maternal deaths and were associated with a severe maternal outcome ratio of 19.4 per 1,000 live births in 2020 [2]. Prospective cohort data from Tanzania also link HDP to elevated maternal and perinatal risk [3]. Early identification of women at risk is therefore an important priority for improving maternal outcomes.

Machine learning (ML) has increasingly been proposed as a means of strengthening maternal risk prediction [4]. By integrating multiple clinical variables and identifying patterns that may not be captured by conventional threshold-based approaches, ML models may support earlier identification and prioritization of high-risk women. However, much of the literature has focused on algorithm development, discrimination, calibration and predictive accuracy. Recent systematic reviews of ML models for preeclampsia report limited evidence on prospective validation or practical deployment detail, particularly for low-resource settings[5, 6]. Comparatively little empirical evidence exists on the health-system, workflow, workforce, and data-system conditions required for ML-enabled risk stratification to function effectively in routine maternal healthcare, particularly in low-resource settings [7].

This gap is particularly important in resource-constrained health systems. Predictive models depend on data generated through routine clinical encounters. The quality, completeness and timeliness of these data are shaped by whether measurements are performed, how they are documented, whether paper records are successfully transformed into digital records, staffing levels, equipment availability, clinical workflows, digital infrastructure and data use practices. An algorithm may demonstrate strong performance in a development dataset yet have limited clinical value if required predictors are not routinely measured, if clinical information does not become timely digital data, if risk outputs arrive too late to influence care, or if frontline providers lack the capacity to respond to additional alerts. These challenges may be particularly pronounced in LMICs, where fragmented documentation systems, human-resource shortages, infrastructure limitations, and variable digital maturity can affect the availability and quality of data required for ML-enabled care [8].

The transition from paper-based to electronic documentation is especially important for ML readiness. In many settings in Africa (including Tanzania), paper and electronic records coexist, especially in primary public health facilities, creating parallel workflows, duplicate documentation requirements, delayed entry and incomplete digital capture [9, 10]. A measurement may therefore be performed during clinical care but never become available for algorithm development. ML models inherit not only biological and clinical patterns, but also the consequences of the documentation and digitization systems that produced their data. The existence of an electronic platform should therefore not be equated with the availability of complete, timely or representative machine-readable data. Drawing on sociotechnical systems theory [11], and calls for responsible, context-appropriate AI implementation in low- and middle-income countries [12], we use the term sociotechnical performance to describe this broader capability: the ability of an ML-enabled intervention to function within real patterns of measurement, documentation, staffing and workflow, rather than being judged solely by statistical performance.

The Mlinde Mama Project was implemented in Geita Region, Tanzania, between 2022 and 2024. The project combined three complementary strategies: redesign of antenatal care through G-ANC; use of Tanzania’s Unified Community System (UCS), to support routine maternal health data capture; and development of ML-enabled risk stratification for HDP. UCS is a national digital health platform designed to support registration, follow-up, referral, and reporting of various health components including maternal and antenatal care services.

G-ANC models have shown feasibility and positive effects on antenatal and postnatal services across various countries in sub-Saharan Africa [13, 14]. In Tanzania, G-ANC was implemented across six public health facilities in two district councils and reached 5,936 pregnant women. The model incorporated clinical assessment, structured health education, peer support and greater participation by women in selected routine measurements and activities [15].

In parallel, the project used routine UCS data to develop a risk stratification ML model for HDP. The best-performing XGBoost model achieved 90.1% accuracy and an area under the receiver operating characteristic curve of 0.95. At the selected high-sensitivity threshold, the model achieved 100% sensitivity but relatively low precision (14%), positioning it primarily as a screening and triage tool rather than a standalone diagnostic system. Despite promising predictive performance, the ML development process was constrained by substantial data-quality challenges, including missingness, duplication, limited longitudinal follow-up, and lacked prospective validation, all of which were identified as important project limitations [16].

These components raise a broader implementation question: what does it take for ML-enabled risk stratification to become useful in routine maternal healthcare? The answer cannot be derived from model performance alone. It requires examining the full pathway through which clinical measurements become digital data, digital data become risk estimates, and risk estimates become clinical action.

This paper examines health systems lessons and implementation readiness considerations based on the Mlinde Mama project experience. Rather than re-evaluating the predictive model, which has been reported separately, we focus here on the health-system conditions surrounding the generation, digitization and potential use of ML-enabled risk information in routine ANC. Guided by the Non-adoption, Abandonment, Scale-up, Spread and Sustainability (NASSS) framework [17] and a study-specific ML development pathway, we examine three linked phases: producing ML-ready data, producing actionable risk information, and converting risk information into care.

## 2. Materials and Methods

### 2.1 Study design

We conducted a retrospective mixed-methods implementation analysis. The analysis integrated participant accounts, project implementation records, empirical characteristics of the routine UCS dataset, and documented experience from development of the ML risk-stratification component. The study followed a convergent triangulation design in which qualitative, quantitative, implementation, and routine data sources were integrated to generate explanatory themes. A sociotechnical perspective guided interpretation: *The ML development process was understood not only as the introduction of a standalone technology, but as an interaction among clinical care, documentation systems, digital technologies, data, healthcare workers, patients, organizational processes and the wider health system*.

### 2.2 Project setting and intervention

The Mlinde Mama Project was implemented in six public health facilities in Geita and Chato District Councils, in Geita region in northwestern Tanzania. This geographical setting is predominantly rural or semi-urban. Participating facilities included district hospitals, health centres and dispensaries. The project integrated G-ANC into routine ANC services. Women of similar gestational age received ANC services in structured group sessions combining clinical assessment, health education, peer interaction and selected self- or peer-supported routine activities[15]. The project also used Tanzania’s UCS to support digital maternal health data capture and development of ML-enabled risk stratification. A total of 5,936 women participated in G-ANC during implementation.

### 2.3 Machine-learning component

The ML component used routine ANC records from the UCS to develop a risk-stratification model for HDP. The technical development and validation have been reported separately. Five models were assessed, with XGBoost selected as the best-performing approach. A total of 337,027 ANC records captured through the UCS between 2023 and 2024 were analysed and subsequently aggregated into 187,438 unique client records for model development. In addition to routinely captured data, clinical and laboratory information from women attending ANC services at the six participating facilities were entered into the UCS during the ML development process to strengthen the dataset used for model training and validation [16].

### 2.4 Data sources

The analysis drew on five complementary forms of evidence: (1) the Mlinde Mama Project Endline Evaluation Report [18], which served as the source document for the focused quotation screening; (2) quantitative exit interviews with 388 pregnant women, including 154 G-ANC and 234 routine ANC participants; (3) qualitative evidence generated through six focus group discussions (FGDs) with pregnant and recently delivered women and 21 key informant interviews (KIIs) with healthcare providers, facility in-charges and district and regional health managers[19]; (4) project implementation records and documented implementation experience relating to the UCS, digital data capture and ML components; and (5) empirical characteristics of the routine UCS dataset observed during ML development, including missingness, duplication, implausible values and temporal sparsity.

The qualitative component was originally designed to examine participants’ experiences of G-ANC, its practicality within routine services, acceptability and implementation challenges, rather than to evaluate ML implementation directly. For the present analysis, we therefore used a focused secondary analytic approach, selecting only qualitative evidence relevant to the broader implementation pathway through which routine maternal healthcare data were generated, documented, digitized, converted into ML-ready data and potentially integrated into clinical workflows. Relevant evidence included participant accounts concerning clinical measurement and self-measurement, documentation and digital data capture, task-sharing, workflow organization, workload, infrastructure and digital equipment, service integration, provider–client relationships, teamwork and sustainability.

### 2.5 Qualitative evidence selection, coding and triangulation

All verbatim participant quotations retained in the final endline evaluation report [16] were screened for relevance to the implementation environment surrounding the digital health and ML components of the Mlinde Mama project. The quotations primarily formed the basis for the broader qualitative study of women’s, healthcare providers’ and health managers’ perspectives on G-ANC implementation in Geita region [19]. Quotations were eligible for inclusion if they provided evidence relating to one or more stages of the implementation pathway: (1) generation of clinical data through care engagement and measurement; (2) documentation, digitization and conversion of routine clinical information into ML-ready data; (3) integration of digital tools and risk information into routine workflows; or (4) broader implementation conditions affecting the digital health intervention, including workload, task-sharing, infrastructure, equipment, teamwork, trust, service integration, governance and sustainability. Quotations concerned primarily with G-ANC experiences that had no clear relevance to data generation, digital transformation, workflow integration or the wider digital health implementation environment were not included in this secondary analysis. Quotation selection was based on relevance to the predefined analytic scope rather than the direction of the finding; both enabling and constraining accounts were eligible for inclusion.

One researcher (AH) conducted the initial screening, selection and coding of eligible quotations. Each quotation was extracted verbatim, distinguished from evaluator narrative, and coded according to participant type, facility or service-delivery context where available, original context in the endline evaluation, implementation-chain phase and proposed analytic theme. A second researcher (AK), who had led the original qualitative study, reviewed the selected quotations, coding and thematic interpretation for consistency with the source data and original study context. Differences in interpretation were resolved through iterative discussion and consensus. Coding was performed manually in Microsoft Excel.

The analysis did not assume that participants were commenting directly on ML or that they had interacted with an ML-generated prediction. Instead, participants’ accounts were treated as evidence about the sociotechnical environment in which data required for ML were produced and in which future ML-generated risk information would need to be interpreted and acted upon.

Qualitative evidence was then triangulated with quantitative evaluation findings, project implementation records and empirical characteristics of the UCS dataset observed during ML development. Triangulation examined convergence, complementarity and tension across evidence sources. For example, participant accounts of self-measurement and task-sharing were considered alongside quantitative evidence on completion of routine measurements; accounts of workload and workflow were interpreted alongside the operational requirements of digital data capture; and evidence concerning documentation and digitization was compared with observed patterns of missingness, duplication, implausible values and temporal sparsity in the ML development dataset. This approach enabled the analysis to trace how routine care became digital data, how digital data became or failed to become-ready, and what organizational conditions would shape the conversion of future ML-generated risk information into clinical action. Conceptual saturation was not an objective of the secondary analysis; instead, thematic sufficiency was assessed through triangulation across data sources.

### 2.6 Analytic framework

We used the NASSS framework [17] to guide secondary analysis and interpretation. NASSS conceptualizes technology-supported healthcare interventions across seven interacting domains: the health condition, technology, value proposition, adopter system, organization, wider institutional context and adaptation over time. NASSS was selected because the study sought to understand not simply whether an ML model achieved adequate predictive performance, but the health-system conditions required for risk stratification to become operationally useful and potentially sustainable within routine ANC.

**Figure 1:**
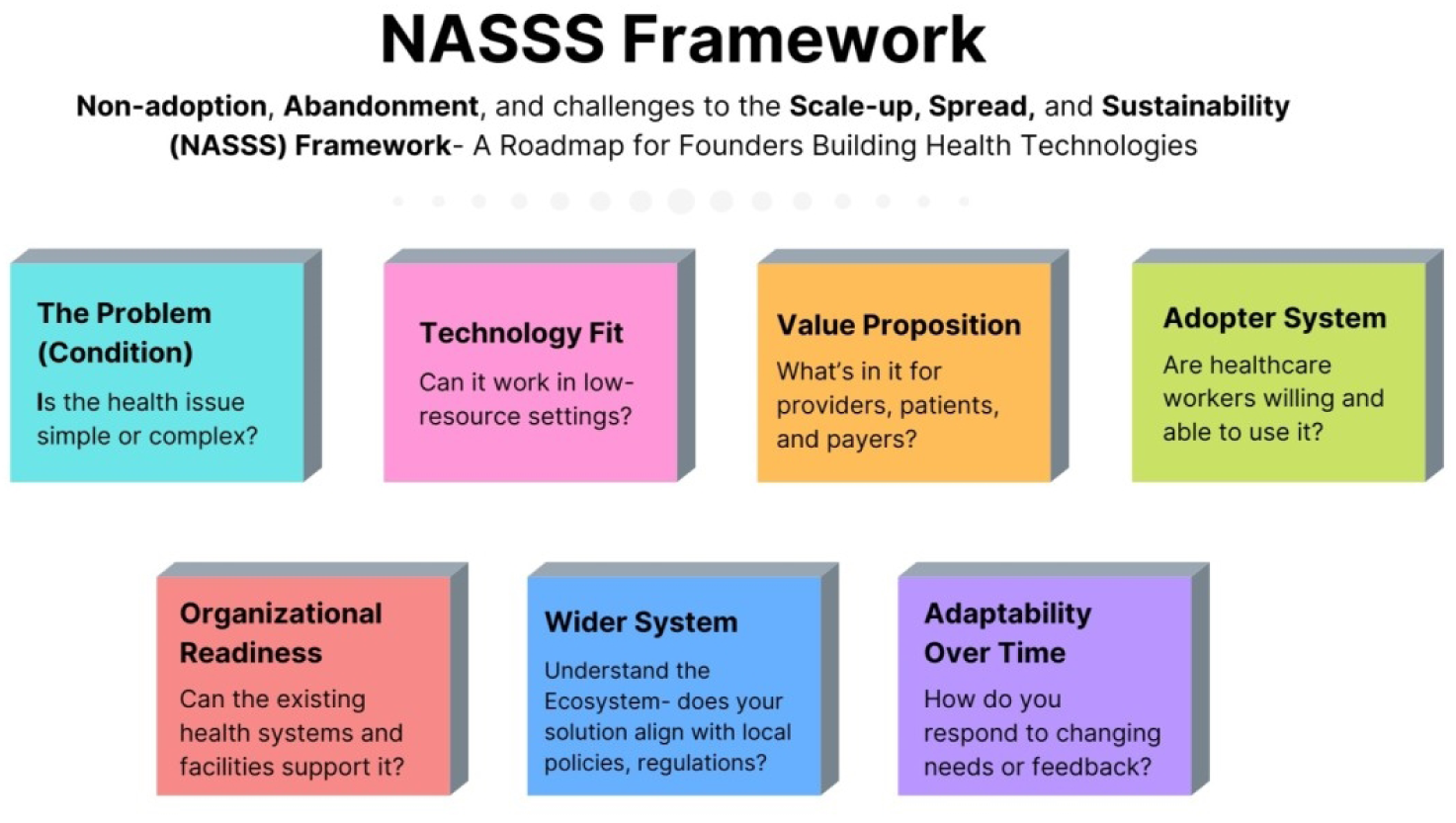
The NASSS Framework. Source: *Adapted from: Greenhalgh et al [17]*

We combined NASSS with a process-oriented ML implementation pathway developed from empirical data. The chain comprised three linked phases. We synthesized the findings into a conceptual model illustrating the pathway (Figure 2). Phase 1, producing ML-ready data, traced the pathway from care engagement through clinical measurement, clinical documentation, the paper-to-digital transition, digitization and digital data quality. Phase 2, converting data into actionable risk information, comprised ML risk estimation, workflow prioritization and clinical confirmation. Phase 3, converting risk information into care, comprised clinical decision-making, targeted action, follow-up or referral and maternal outcome. Infrastructure, workforce, trust and governance were treated as cross-cutting conditions influencing all three phases.

**Figure 2.**
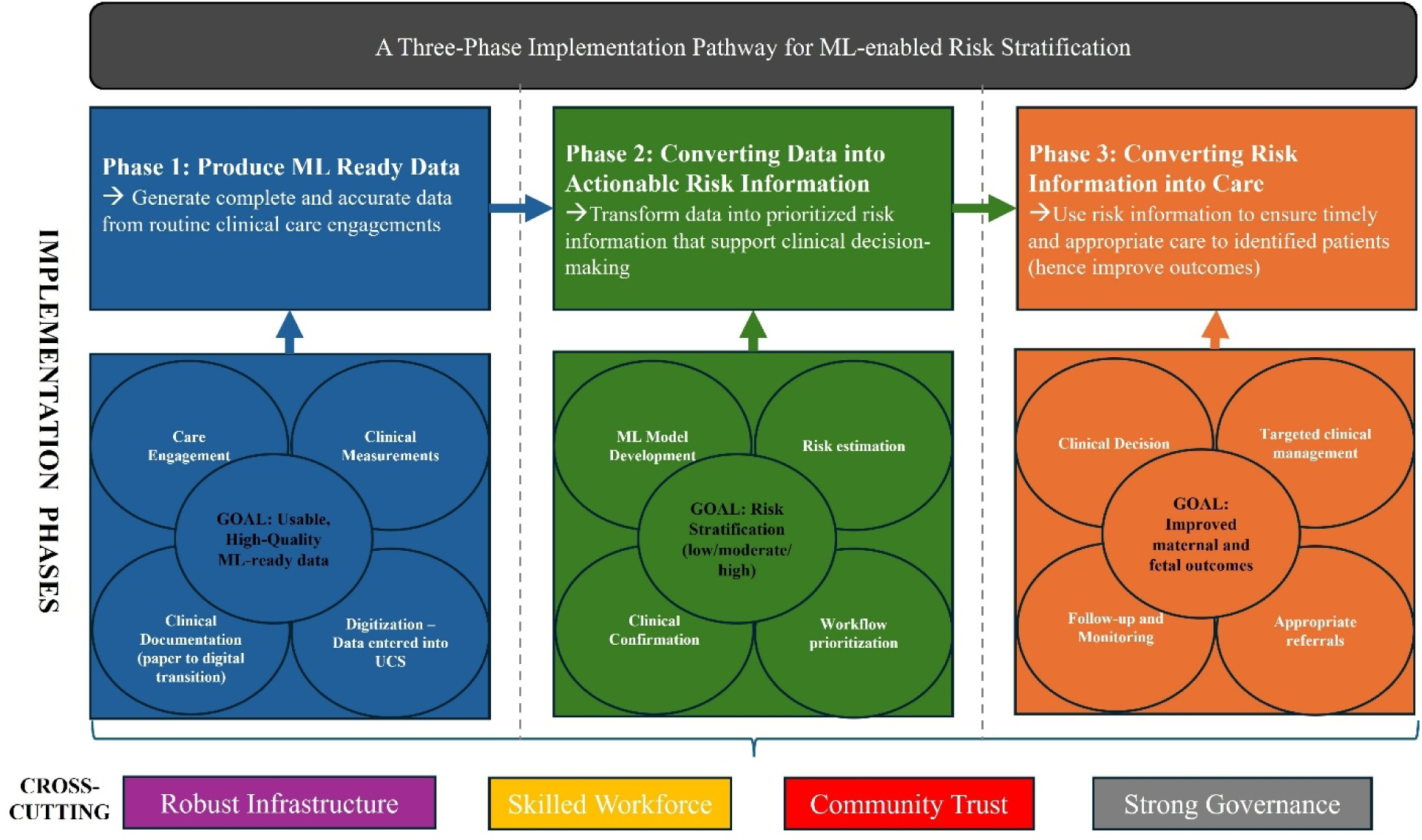
A three-phase implementation pathway for ML-enabled maternal risk stratification. Phase 1 represents the production of ML-ready data from routine clinical care and highlights the paper-to-digital transition gap, where parallel systems, duplicate entry, delayed entry and incomplete capture may create divergence between clinical and digital reality. Phase 2 represents the conversion of digital data into actionable risk information through ML risk estimation, prioritization and clinical confirmation. Phase 3 represents the conversion of risk information into clinical decisions, targeted action and follow-up. Infrastructure, workforce, trust and governance influence all phases, while feedback from care and outcomes supports learning and adaptation over time.

The analysis was primarily abductive. Participant quotations and other project evidence were first examined according to their original meaning and implementation context. Emerging themes were then compared with NASSS domains and stages of the ML implementation chain. This allowed the analysis to remain grounded in project evidence while using theory to interpret interactions among clinical, technological and organizational conditions. The three-phase implementation pathway was used to organize where implementation challenges arose along the journey from routine care to clinical action, while NASSS was used to interpret the wider technological, organizational and contextual factors shaping each phase.

### 2.7 Ethical considerations

The endline evaluation was undertaken as part of the approved implementation research activities of the Mlinde Mama Project. Participants provided informed consent before interview. This secondary analysis uses de-identified participant quotations and aggregated implementation data. The study received ethical approval from the National Institute for Medical Research (NIMR) through the National Health Research Ethics Committee (NIMR/HQ/R.8a/Vol.IX/4194; 23 January 2023). The approved protocol permits secondary analysis of de-identified project data.

## 3. Results

Seven interconnected themes were identified across the three phases of the ML implementation pathway (Figure 2). Themes 1–3 concerned the production of ML-ready data; Theme 4 concerned the conversion of data into actionable risk information; and Themes 5–7 described cross-cutting organizational and relational conditions shaping implementation across the pathway. An overview of the seven themes, their evidence sources and interpretive contributions is provided in Table 1.

**Table 1.** Overview of themes, evidence sources, implementation phases and interpretive contributions, mapped to the NASSS Domains.

|  | Theme | Implementation-chain phase | Primary evidence | Main NASSS domains | Interpretive contribution |
| --- | --- | --- | --- | --- | --- |
| 1. | ML readiness began before the algorithm development | Producing ML-ready data | Participant quotations on blood pressure and weight measurement | Condition, Technology; Adopter system | Prediction depends on reliable generation of clinical measurements. |
| 2. | Redistributing measurement and data generation | Producing ML-ready data | Participant quotations on self/peer activities and task-sharing | Value proposition<br>Adopter system;<br>Organization | Who performs routine work shapes scalability and data availability. |
| 3. | From clinical reality to digital reality | Producing ML-ready data | Triangulation of evaluation findings, implementation experience and UCS dataset characteristics | Technology; Adopter system; Organization;<br>Wider context | Clinical reality and digital reality may diverge before data reach the model. |
| 4. | Workflow determined clinical value | Converting data into actionable risk information | Provider quotation plus ML operating characteristics | Technology, Adopter System; Organization | A prediction becomes useful only through a feasible confirmation and action pathway. |
| 5. | The efficiency paradox | Cross-cutting across all phases | Positive and negative provider cases | Value proposition<br>Adopter system;<br>Organization | Technology may save some work while creating new tasks and alert burden. |
| 6. | The hidden infrastructure beneath AI | Cross-cutting across all phases | Provider quotations on equipment and phones | Technology;<br>Organization; Wider context | Basic material conditions are components of the functioning ML system. |
| 7. | Trust, teamwork and ownership | Cross-cutting across all phases, with relevance to Phase 3 | Participant quotations on relationships, teamwork and demand | Adopter system;<br>Organization; Wider context, Adaptation over time | Risk information becomes action through relationships and collective implementation. |

### 3.1 Theme 1: ML readiness began before the algorithm: generating the clinical measurements

The development of the ML model depended on routinely generated clinical data. The project experience demonstrated, however, that such data could not be treated simply as an existing technical resource. Data were produced through interactions among pregnant women, healthcare workers, measurement devices and clinical workflows.

Within G-ANC, women became more actively involved in selected routine measurements. A pregnant woman described this process:

> *"Many pregnant women gained the ability to measure their own weight and blood pressure and encouraged others to join the groups."* Pregnant woman, District Hospital 1.

A healthcare provider similarly observed:

> *"They learned various things, such as measuring their own blood pressure and teaching each other."*

Healthcare provider, Health Centre 1.

These accounts suggest that ML readiness began with the reliable production of clinical measurements. Blood pressure and other physiological measurements were not merely variables in a dataset; they were products of specific clinical and social practices. This was part1icularly relevant because blood pressure was central to the HDP risk-stratification model. The model could only identify risk when relevant measurements were performed, documented and made available in usable digital form.

This theme primarily relates to the *Condition, Technology*, and *Organization* domains of the NASSS framework. It demonstrates that ML readiness begins with the reliable generation of routine clinical measurements rather than with algorithm development itself. The quality of risk prediction therefore depends on organizational capacity to consistently produce accurate clinical information as part of routine antenatal care.

### 3.2 Theme 2: Redistributing the work of measurement and data generation

The G-ANC model redistributed some aspects of routine care from an exclusively provider-led process towards greater participation by pregnant women and peer groups. This did not replace professional clinical assessment. Rather, it created opportunities for routine information to be generated earlier and for women to become more engaged in their care.

One provider described the resulting workflow:

> *"When a pregnant woman comes, she has already been tested, which reduces the time she spends with healthcare provider at the hospital."* District Hospital 2.

Another explained:

> *"It was easier to serve [G-ANC members] because they handled many things themselves, allowing us to spend less time with them."* Healthcare provider, District Hospital 1.

Participants consistently described task-sharing as reducing provider workload and facilitating service delivery. Distributed data generation may strengthen the upstream data pipeline while preserving clinician attention for tasks requiring professional judgement.

This theme primarily reflects the *Value proposition, Adopter System* and *Organization* domains of the NASSS framework. The redistribution of measurement and documentation activities altered the roles of women, healthcare providers and peer groups, illustrating that successful digital health implementation depends on how new responsibilities are integrated into existing clinical workflows and accepted by those delivering and receiving care.

### 3.3 Theme 3: From clinical reality to digital reality: the paper-to-digital transition shaped ML-ready data

The project experience demonstrated that performing a clinical task did not necessarily mean that the resulting information became available for digital analysis. Between measurement at the point of care and inclusion in the ML dataset lay an intermediate process of documentation, digital entry, data transmission and record linkage.

The Mlinde Mama intervention was implemented within a health system transitioning from predominantly paper-based documentation towards greater use of digital systems. Routine ANC information could therefore exist across paper registers (such as the facility ANC registers, labour and delivery register and the cohort tracking register), patient-held records such as the RCH Card and the UCS. The coexistence of documentation systems meant that digitally available information did not necessarily represent the complete set of clinical activities performed during an ANC encounter.

This distinction became visible when evaluation findings were compared with the data available for ML development. In the endline evaluation, all interviewed women reported having their blood pressure measured during the current or a previous ANC visit. Weight measurement was reported by 98.7% of G-ANC participants and 93.2% of women receiving routine ANC[18]. Yet the companion ML study encountered substantial incompleteness and temporal sparsity in routinely captured digital records[16].

The ML development process required extensive transformation of routine UCS data, including removal of duplicate entries, correction of implausible physiological values and management of missing observations. Multiple visits also had to be collapsed into single patient-level records because the longitudinal digital record was sparse: the median number of recorded visits per client was one, and only 43% of clients had two or more digitally recorded visits[18].

Taken together, these findings revealed a gap between clinical reality and digital reality. A measurement could be performed but not documented; documented on paper but not entered digitally; entered after a delay; duplicated across systems; or captured in a form requiring substantial cleaning before computational use. Consequently, the data available to the ML model represented the subset of clinical information that successfully passed through the documentation and digitization pathway.

The triangulated evidence identified four plausible mechanisms through which information could be lost or distorted during this transition: parallel documentation, duplicate entry, delayed entry and incomplete digital capture. Data quality was therefore not solely a property of the analytic dataset; it was produced by the wider care and documentation system.

The "paper-to-digital" transition represents a point of convergence across multiple NASSS domains, including the *Technology* (digital information systems), *Adopter System* (healthcare providers responsible for documenting and entering clinical information), *Organization* (workflow design, documentation practices and system readiness), and the *Wider Context* (national digital health policies, governance and reporting requirements). Consequently, failures in digitization should not be viewed solely as technical problems but as manifestations of broader sociotechnical challenges within the health system.

### 3.4 Theme 4: From prediction to action: workflow determined clinical value

The technical ML model was designed primarily as a risk-stratification and triage tool. Its intended value was therefore not simply to generate a risk prediction but to help determine which women required additional attention.

Providers valued interventions that allowed information to be available before or early in the clinical encounter, as noted above (Section 3.2): *"When a pregnant woman comes, she has already been tested, which reduces the time she spends with health care provider at the hospital."*

This experience parallels the proposed role of ML-enabled triage. Given the model’s operating characteristics, a future high-risk flag would require clinical confirmation rather than automatically triggering specialist referral or treatment and should instead be prioritized for confirmatory assessment.

The model’s high sensitivity and low precision make workflow design especially important. A high-risk flag should not automatically trigger specialist referral or treatment. Instead, it could trigger a structured next step such as repeat blood pressure measurement, review of proteinuria, targeted clinical assessment or expedited provider review. The output of an algorithm is therefore not the end of the intervention; it is the beginning of a workflow. This finding primarily relates to the *Technology, Adopter System*, and *Organization* domains of the NASSS framework. The clinical value of ML depended not on the prediction itself but on how risk information was interpreted, confirmed and incorporated into routine workflows. Algorithm outputs therefore become useful only when embedded within clearly defined clinical pathways and supported by organizational processes.

### 3.5 Theme 5: The efficiency paradox: technology could save and create work

Providers described ways in which the intervention reduced routine workload. One participant reported:

> *"The tools provided, like the battery machine and weight scale, reduced our workload."* Healthcare provider, District Hospital 1.

Task-sharing also allowed providers to concentrate on activities requiring greater clinical input. However, the evaluation identified an important negative case:

> *"In one day, we would have more than six groups, and we spent a lot of time serving them due to the shortage of staff."* Healthcare provider, District Hospital 2.

This apparent contradiction is important for ML implementation. Technology may reduce the time required for some activities while simultaneously increasing demand, generating new tasks or identifying more patients who require follow-up. This is particularly relevant to a high-sensitivity model with relatively low precision because false-positive alerts may generate additional confirmatory work. This is quite important in the context of the NASSS framework. Efficiency should therefore be assessed across the full pathway—from measurement to risk flag, confirmation and clinical action. This theme primarily reflects the *Value proposition*, *Adopter System, Organization* and *Adaptation over time* domains of the NASSS framework. Technologies perceived as helping to solve routine work challenges (i.e. are valuable), are more likely to be adopted. Conversely, technologies that are perceived to increase workload or introduce additional clinical tasks are less likely to be adopted and sustained [20].

### 3.6 Theme 6: The hidden infrastructure beneath AI

One of the clearest findings was the dependence of digital innovation on basic physical resources. Providers appreciated equipment that supported routine measurement, as noted above (Section 3.5): "The tools provided, like the battery machine and weight scale, reduced our workload."

At the same time, a provider’s request for continued support revealed the material realities underlying digital health implementation:

> *"Additionally, I request the project to assist us with work phones, bags to store the phones, and rain gear to protect the equipment when it rains."* Healthcare provider, District Hospital 1.

A healthcare manager said:

> *“…we don’t have money to print maternal booklets and to replace rechargeable batteries for BP machines.” —* District Coordinator, District 2.

An advanced predictive model may depend on whether a frontline worker has a functioning phone or tablet, whether devices are protected during travel and rain, whether batteries can be charged, whether measurement equipment, and whether tools for data documentation are available. These are not peripheral implementation issues; they are components of the functioning ML system. This theme mainly reflects the *Technology, Organization,* and *Wider Context* domains of the NASSS framework. Reliable AI implementation depends on basic infrastructure, including functional equipment, power supply, digital devices, documentation tools and sustainable financing. These seemingly simple resources are integral components of the digital health ecosystem rather than peripheral implementation considerations.

### 3.7 Theme 7: From adoption to sustainability: trust, teamwork and ownership

The sustainability of the intervention was linked not only to technology and equipment but also to relationships among patients, providers and teams.

A pregnant woman described improved communication:

> *"There has been a strong relationship between the service provider and the client, where they feel free to express their challenges."* Pregnant woman, District Hospital 1.

A healthcare provider emphasized teamwork:

> *"The key to success of the project was teamwork, which made the work easier."* Healthcare provider, Health Centre 2.

Demand from women was also evident:

> *"We request that the project continues, as the mothers have been highly motivated to learn and take care of themselves."* Healthcare provider, Health Center 2.

These findings suggest that ML-enabled care will be introduced into an existing network of relationships. Whether a woman accepts further evaluation, communicates symptoms or returns for follow-up may depend on trust and perceived quality of care. Similarly, risk stratification may redistribute work across nurses, clinicians, community health workers and digital systems. Teamwork is therefore part of the mechanism through which risk information becomes clinical action. This theme primarily reflects the *Adopter System, Organizatio*n, *Wider context* and *Adaptation over Time* domains of the NASSS framework. Trust between women and healthcare providers, teamwork across professional groups and local ownership emerged as essential conditions for sustaining implementation. Long-term integration of ML-enabled care will therefore depend not only on technological performance but also on the capacity of health systems to maintain collaborative relationships, adapt workflows and support continuous learning.

## 4. Discussion

### 4.1 Principal findings

The central finding is that ML implementation began before the algorithm and extended beyond prediction. Seven interrelated lessons mapped onto three phases of the implementation pathway. Producing ML-ready data required reliable measurement, appropriate task-sharing and successful movement of information from clinical documentation into digital systems. Producing actionable risk information required workflow redesign and feasible confirmation pathways. Converting risk information into sustainable care depended on workload, infrastructure, trust, teamwork and organizational capacity.

This framing extended beyond conventional assessments of ML readiness. Data quality was a care-delivery and documentation issue as much as a technical issue. The paper-to-digital transition emerged as a hidden stage in the ML pipeline. Infrastructure and relationships were not peripheral facilitators; they were part of the functioning sociotechnical system.

The seven themes intersected across multiple NASSS domains rather than mapping to single domains in a one-to-one manner. The technology, adopter and organizational domains were particularly prominent, while the wider context and adaptation-over-time domains shaped the potential for scale and sustainability. The relationship between the Mlinde Mama findings and the seven NASSS domains is summarized in Table 2.

**Table 2.** Mapping of NASSS domains to the Mlinde Mama findings.

| NASSS domain | Application to Mlinde Mama | Aligned themes |
| --- | --- | --- |
| Condition | Clinical complexity and need for timely identification of women at risk of HDP within high-volume ANC services | Theme 1 |
| Technology | Measurement devices, UCS, paper-to-digital transition, data quality and ML model | Themes 1, 3, 4 and 6 |
| Value proposition | Earlier risk identification and better allocation of scarce provider attention | Themes 2 and 5 |
| Adopter system | Pregnant women, providers, CHWs and facility teams | Themes 2,3,4,5 and 7 |
| Organization | Documentation practices, paper-to-digital transition, staffing, task-sharing, workflow redesign, sustainability | Across all themes (1-7) |
| Wider context | National digital-health architecture (UCS), governance, policy and institutional arrangements. | Themes 3, 6 and 7 |
| Adaptation over time | Evolution of data systems, workflows and sustainability mechanisms | Themes 5 and 7 |

### 4.2 From predictive performance to sociotechnical performance

The technical ML study demonstrated that routine UCS data could support high-sensitivity risk stratification for HDP. However, conventional metrics describe only one dimension of performance. For routine implementation, a model must also demonstrate sociotechnical performance: *the ability to function within actual patterns of measurement, documentation, digitization, staffing, patient flow and clinical decision-making*.

A model with excellent discrimination may have little clinical effect if required inputs are missing or delayed. Conversely, a model with high sensitivity may create substantial operational burden if every alert leads directly to specialist referral. This echoes broader sociotechnical systems accounts of AI in healthcare delivery, which argue that the entire work system, not the algorithm alone, determines whether AI improves patient safety and reduces clinician burden [11], and aligns with calls for AI implementation in LMICs to be treated as an exercise in responsible, sustainable and inclusive health-system innovation rather than a purely technical deployment [12, 21]. Staffing constraints identified in the qualitative analysis further illustrate how workforce capacity may shape ML implementation and reinforce the potential role of appropriately designed task-sharing [22, 23]. In high-volume facilities, requiring clinicians to perform every routine measurement, enter every data point and interpret every risk signal may limit scalability. This signifies the workload the clinician must undertake in producing high-quality data. Therefore, structured task-sharing has the potential to reduce the workload associated with selected routine activities. Thus, the Mlinde Mama experience supports a broader definition of ML performance that includes data availability, digital representativeness, workflow fit, actionability, workload consequences, user trust, equity and sustainability.

### 4.3 The paper-to-digital transition as a hidden stage in the ML pipeline

The results revealed that the transition from clinical care to machine-readable data was not seamless. ML models are often described as being developed from "routine health data," implying that information generated during care is automatically available for computational analysis. In practice, clinical reality and digital reality are not equivalent.

The coexistence of paper and electronic systems created several opportunities for information loss or distortion. A measurement may be have been performed but not documented, remain only on paper, be entered after a delay, be duplicated across systems, or require substantial cleaning before analysis. This finding is consistent with broader evidence on routine health information systems in sub-Saharan Africa: facility-reported DHIS2 data have been found incomplete at least 40% of the time in some settings, with 10–60% of clinically documented events never captured in the digital system [24], and persistent completeness, timeliness and consistency gaps have been documented across DHIS2 implementations more broadly, including in Tanzania [25]. Further evidence from other health systems assessing integration aspects of digital data systems similarly identifies parallel documentation, duplicate work, incomplete capture and workflow disruption as persistent challenges [9, 10, 26, 27].

This has important implications for interpreting missing data in ML development. Missingness should not be understood solely as a statistical property of the final dataset; it may be a trace of the underlying health system. Missing values can reflect unavailable equipment, stock-outs, selective testing, staff workload, parallel documentation, poor connectivity, delayed entry or system-design weaknesses. Imputation may improve an analytic dataset but cannot correct the upstream implementation conditions that produced the missingness.

The distinction also has implications for algorithmic equity. If some facilities, patient groups or encounters are less likely to be digitally documented, they may become systematically underrepresented in the data used to develop and operate an ML model. The algorithm may then reproduce patterns of digital visibility rather than the true distribution of clinical need. Although the present study did not directly evaluate algorithmic bias, these findings highlight an important ethical consideration for AI implementation in resource-limited settings: responsible AI requires attention not only to model performance, but also to who is represented in the underlying digital data and who may remain invisible [28].

ML readiness should therefore not be equated with the existence of an electronic platform. The more relevant question is whether clinically meaningful information is captured completely, accurately, consistently and sufficiently close to the point of care to support timely risk stratification and action. Strengthening the paper-to-digital transition may be as important as improving the model itself. The “paper-to-digital” transition emerged as a critical point of convergence across the NASSS framework. It simultaneously reflects the capabilities of the technology, the practices of healthcare providers, the readiness of the organization, and the influence of the wider policy and governance environment. As such, failures in digitization should not be viewed solely as technical problems, but as manifestations of broader sociotechnical challenges within the health system.

### 4.4 The algorithm as part of a clinical pathway

The findings reinforce the argument that ML outputs should not be treated as clinical endpoints. The high-sensitivity model was best suited to triage rather than diagnosis. A plausible future workflow is:

Routine measurement → Digital capture → Automated risk stratification → Targeted confirmation → Clinician review → Referral or follow-up.

This is more realistic than either fully automated decision-making or adding ML alerts on top of unchanged clinical workflows. Each risk category should correspond to a feasible clinical action, and the consequences of false-positive alerts should be explicitly designed and measured. In this sense, the output of the model is valuable only when the downstream pathway can convert risk information into timely care.

### 4.5 The hidden infrastructure beneath AI

The provider’s request for phones, bags and rain protection illustrates a central implementation reality: AI in resource-constrained settings is inseparable from the devices, power systems, connectivity, measurement equipment and environmental conditions through which data are generated and transmitted. Studies in both developed and resource-constrained settings highlight system-level challenges that must be solved before the technology can efficiently produce the intended outcomes[29, 30].

This has implications for investment decisions. Funding an algorithm without financing the infrastructure required to produce its inputs and act on its outputs risks creating a technically impressive but operationally fragile intervention.

### 4.6 Implications for implementation and scale-up

Our findings suggest that implementation and scale-up of ML-enabled maternal risk stratification should proceed through a phased health-system approach rather than through deployment of the algorithm alone:

- **Establish readiness to produce ML-ready data:** Before model deployment, implementers should determine whether the health system can reliably generate the data on which the model depends. This requires mapping the pathway from clinical measurement to digital availability; identifying which predictors are routinely measured; quantifying missingness and implausible values by facility and patient group; and identifying where parallel paper and digital systems create duplicate, delayed or incomplete data capture. Major upstream data gaps should be addressed before algorithmic outputs are used for clinical prioritization.
- **Make the paper-to-digital transition an explicit implementation workstream.** In settings where paper-based data systems are still predominant, programs should define where each data element is first recorded, who is responsible for entering it digitally, when digital entry should occur, and how discrepancies between paper and electronic records will be resolved. Routine monitoring should include indicators of digital completeness, timeliness, duplication and longitudinal continuity rather than relying only on aggregate measures of data quality.
- **Design the pathway from risk prediction to clinical action.** ML outputs should be linked to predefined clinical responses before deployment. For each risk category, implementers should specify who receives the alert, the expected response time, the confirmatory assessment required, who makes the final clinical decision, and what follow-up or referral pathway is available. High-risk classifications should trigger structured clinical review rather than automated treatment or referral.
- **Measure the work created as well as the work saved.** Prospective implementation should assess workload across the entire pathway. Evaluation should capture not only time saved through automation or task-sharing, but also additional work generated by digital documentation, alert review, repeat measurements, clinical confirmation and false-positive risk classifications. Staffing and workflows should be adapted when alert volume exceeds the capacity of the service to respond safely.
- **Define readiness for expansion.** Decisions to introduce the intervention into additional facilities should be based on minimum readiness criteria, including the availability and functionality of essential measurement equipment, access to appropriate digital devices and connectivity, staff capacity, data-governance arrangements, referral capacity and mechanisms for clinical oversight. Expansion should depend not simply on whether a facility can run the technology, but on whether it can reliably generate the required data and act safely on the resulting risk information.
- **Scale through learning and adaptation.** Because the complete ML-enabled clinical pathway has not yet been prospectively evaluated, the immediate priority is not widespread deployment but phased implementation, adaptation and evaluation. Initial implementation should involve a limited number of representative facilities and prospectively monitor data completeness, digital representativeness, alert burden, workflow effects, equity, provider and patient experience, costs and clinical outcomes. Findings should be used to refine the intervention before wider expansion. Any subsequent progression to scale should follow a deliberate strategy that considers the innovation itself, the organizations expected to adopt it, the wider health-system environment, resource requirements and mechanisms for institutionalization [31].

### 4.7 Strengths and limitations

This analysis draws on implementation of digital health, service-delivery redesign and ML development within a real public health system rather than a controlled research environment. It links technical model development with frontline experiences of measurement, workflow, workload and sustainability, and triangulates qualitative, quantitative, implementation and routine-data evidence.

Several limitations require emphasis. Most importantly, the qualitative interviews were not designed specifically to examine perceptions of ML or artificial intelligence, and participants did not directly discuss the algorithm. The present analysis therefore interprets their experiences as evidence about the clinical and organizational environment into which ML-enabled risk stratification would be introduced.

Second, the qualitative re-analysis was based primarily on quotations retained in the final evaluation report rather than complete interview transcripts. This limits depth and may introduce selection bias. Although quotation selection and coding were reviewed by a researcher who led the original qualitative study, the analysis could not recover contextual detail that was not retained in the report. Third, the paper-to-digital transition theme relies on triangulation of evaluation findings, implementation experience and empirical characteristics of the UCS dataset rather than direct participant testimony. This distinction should remain explicit.

Fourth, the ML model was developed and validated retrospectively because the Mlinde Mama project came to an end before that stage could be completed. The full ML-enabled workflow was not prospectively evaluated at the point of care. Claims regarding clinical efficiency should therefore be treated as implementation hypotheses requiring prospective testing. Additionally, this study examines implementation readiness and health-system conditions for future ML deployment rather than evaluating a deployed ML intervention. Finally, findings emerged from six facilities in one region of Tanzania and may not capture the full diversity of infrastructure, staffing and digital-system conditions nationally.

## Conclusion

The Mlinde Mama experience demonstrates that preparing ML for maternal healthcare in Africa and other low-resource settings involves far more than inserting an algorithm into clinical care. The pathway begins upstream of the algorithm, with care engagement, clinical measurement, documentation and the successful transformation of clinical information into usable digital data. It extends beyond prediction to workflow prioritization, clinical confirmation, communication, clinical judgement and appropriate action.

The central lesson is that the algorithm was only one component of the intervention. For ML-enabled maternal risk stratification to improve care in resource-constrained settings, investments must extend beyond model development to the data-generating practices, paper-to-digital transition, clinical workflows, equipment, workforce, relationships and governance structures that enable predictions to be translated into clinical action.

The key lesson learnt is that, implementing AI in routine maternal healthcare is fundamentally a health-system challenge rather than solely a technical one. Viewed through the NASSS framework, ML-enabled maternal risk stratification is best understood as a sociotechnical intervention whose success depends on coordinated attention to technology, adopters, organizational readiness, infrastructure and the wider policy and governance environment. Future AI implementation efforts in low-resource settings should therefore invest as much in strengthening routine health systems as in developing predictive algorithms. Future research should prospectively evaluate ML-enabled triage within routine antenatal care, measuring not only predictive performance but also implementation outcomes, including digital data completeness, workflow integration, provider workload, patient experience, equity, costs and clinical outcomes, to better understand how AI can be effectively embedded within routine health systems.

## What this study adds

### What is already known

Machine learning can identify maternal risk using routinely collected clinical data, but most studies emphasize predictive performance rather than the real-world pathway through which clinical information becomes usable data and then clinical action.

### What this study adds

The Mlinde Mama experience shows that ML readiness begins upstream of the algorithm. Reliable measurement, task-sharing and the paper-to-digital transition determine which clinical information becomes ML-ready data; workflow design then determines whether risk predictions become actionable.

### Implications for policy and practice

ML-enabled maternal risk stratification should be implemented as a sociotechnical intervention spanning measurement, documentation, digitization, data quality, prediction, confirmation and clinical action. Scale-up should combine algorithms with reduction of parallel documentation, point-of-care digital capture, workflow redesign, workforce investment and basic infrastructure.

## Data Availability

The de-identified quantitative dataset and accompanying data dictionary are publicly available in Harvard Dataverse [30] (DOI: 10.7910/DVN/RLAJMC). Qualitative quotations analysed in this study were drawn from the published Mlinde Mama Endline Evaluation Report [16]. Additional project implementation records and routine UCS data are subject to applicable institutional and government data-governance requirements.

## Acknowledgements

The Mlinde Mama Project was implemented by Prime Health Initiative Tanzania in partnership with the Tanzania Ministry of Health, the Prime Minister’s Office – Regional Administration and Local Government (PMO-RALG), local government authorities and the Geita Regional Health Management Team. We thank the participating women, healthcare providers, facility leaders, community health workers and government partners whose collaboration made the Mlinde Mama Project possible. We are particularly grateful to the women attending antenatal care services, healthcare providers and health managers who participated in the qualitative assessment and shared their experiences and perspectives, which were invaluable to understanding the health-system conditions shaping implementation of digital health and ML-enabled maternal risk stratification.

## Data Reporting

The de-identified quantitative dataset and accompanying data dictionary are publicly available in Harvard Dataverse [32] (DOI: 10.7910/DVN/RLAJMC). Qualitative quotations analysed in this study were drawn from the published Mlinde Mama Endline Evaluation Report [16]. Additional project implementation records and routine UCS data are subject to applicable institutional and government data-governance requirements.

## Financial Disclosure Statement

The Mlinde Mama Project was supported by the Gates Foundation through the Grand Challenges Global Call-to-Action: Digital Health Services (Investment ID INV-046249). The funder had no role in the design of this secondary analysis, interpretation of findings, decision to publish or preparation of the manuscript. The views expressed are those of the authors and do not necessarily reflect those of the Gates Foundation.

## Competing interests

The authors declare no competing interests.

## Author contributions

AH: Conceptualization, Methodology, Formal analysis, Visualization, Writing – original draft, Writing – review & editing. AK: Validation, Investigation, Writing – review & editing. EE: Formal analysis, Data curation, Writing – review & editing. IL and YK: Methodology, Software, Formal analysis, Data curation, Writing – review & editing. HM, CM, HA, FP, PM, OS, PS, NK: Investigation, Resources, Project administration, Supervision, and Writing – review & editing. All authors reviewed and approved the final manuscript.

## Notes

### Competing Interest Statement

The authors have declared no competing interest.

### Author Declarations

The endline evaluation was undertaken as part of the approved implementation research activities of the Mlinde Mama Project. Participants provided consent before interview. This secondary analysis uses de-identified participant quotations and aggregated implementation data. The study received ethical approval from the National Institute for Medical Research (NIMR) through the National Health Research Ethics Committee (NIMR/HQ/R.8a/Vol.IX/4194 23 January 2023).

